# Benchmarking Vision Encoders for Survival Analysis using Histopathological Images

**DOI:** 10.1101/2024.08.23.24312362

**Authors:** Asad Nizami, Arita Halder

## Abstract

Cancer is a complex disease characterized by the uncontrolled growth of abnormal cells in the body but can be prevented and even cured when detected early. Advanced medical imaging has introduced Whole Slide Images (WSIs). When combined with deep learning techniques, it can be used to extract meaningful features. These features are useful for various tasks such as classification and segmentation. There have been numerous studies involving the use of WSIs for survival analysis. Hence, it is crucial to determine their effectiveness for specific use cases. In this paper, we compared three publicly available vision encoders-UNI, Phikon and ResNet18 which are trained on millions of histopathological images, to generate feature embedding for survival analysis. WSIs cannot be fed directly to a network due to their size. We have divided them into 256 × 256 pixels patches and used a vision encoder to get feature embeddings. These embeddings were passed into an aggregator function to get representation at the WSI level which was then passed to a Long Short Term Memory (LSTM) based risk prediction head for survival analysis. Using breast cancer data from The Cancer Genome Atlas Program (TCGA) and k-fold cross-validation, we demonstrated that transformer-based models are more effective in survival analysis and achieved better C-index on average than ResNet-based architecture. The code^1^ for this study will be made available.

## 1 Introduction

In recent years, cancer has become a major global health concern, resulting in nearly 10 million deaths each year. This makes cancer the leading cause of death after cardiovascular disease [1]. By 2040, it is expected that 29.9 million new cancer cases will be diagnosed each year, with 15.3 million cancer-related deaths^2^. The type of cancer contributing to these statistics varies by gender and region. The most common types of cancer include breast, lung, colon, and rectal cancer. Cancer is treatable in many cases, especially when they are detected early.

Histopathological images like Whole Slide Images (WSIs) have emerged as powerful tools in cancer detection and early prognosis. Using advanced imaging and computational tools, WSIs can enhance diagnosis and treatment planning for cancer. Pathological images obtained from Whole Slide Images (WSI) provide detailed visual representations of tissue. Deep learning methods [2, 3, 4, 5] have shown great potential in extracting meaningful features for different tasks such as the classification, survival analysis and segmentation of cancerous cells. This way they are effective in diagnosing cancer and have aided in the early prognosis. However, working with WSIs has some challenges of its own. They have very high resolution-100,000 × 100,000 pixels and are often several gigabytes in size. Analyzing these files requires significant computational resources.

The advanced digital scanner has aided in scanning tissue samples and shifted the workflow bottleneck from image acquisition to image annotation. Techniques like self-supervised learning (SSL) [6, 7] and weakly-supervised learning [8, 9, 10] are useful for training a neural network on unannotated images, which can then be fine-tuned on annotated datasets for specific downstream tasks. These approaches enable the use of large amounts of unannotated datasets, which are often more readily available than annotated ones. The model can learn useful representation by training a neural network on these datasets, reducing the amount of annotated data required for fine-tuning.

Survival analysis is a branch of statistics that deals with the analysis of time-to-event data. It is particularly useful in medical research as it can help predict patient outcomes over time. Using covariates which can be extracted from WSIs, one can predict the relative risk scores of the data samples. Multimodal studies [11, 12, 13] have contributed significantly to medical research. Integrating various types of inputs like histopathological images and clinical data of patients has enhanced predictive modelling and clinical decision-making. However, collecting clinical data requires more manual tasks as compared to acquiring tissue samples and generating digital images for analysis. In this study, we compare the efficiency of several vision encoders for survival analysis by only using Whole slide images from the breast cancer repository (BRCA) of The Cancer Genome Atlas Program (TCGA). Since whole slide images (WSIs) are too large to fit directly in a neural network, many studies use Multiple Instance Learning (MIL) to generate feature embedding at the WSI level. This encoded data can then be fed directly to a network for various tasks.

Recently we have many studies done to get a representation of WSIs. Therefore, it is necessary to compare them to determine which works best for specific tasks. In particular, we compared three vision encoders for their application in survival analysis. In summary, our contributions are as follows.

1. Review the recent literature to understand the prior work.
2. Training a network, using the vision encoders for feature embeddings. The goal of the network is to predict relative risk scores among the data samples.
3. Comparing the performance of three pretrained encoders for survival analysis.

The following section introduces an overview of the state-of-the-art literature to study the application of histopathological images and deep learning. Section 3, introduces the relevant technologies, concepts as well as the experimental setup used in this research. The results of the experiments are discussed in Section 4 followed by a conclusion in Section 5.

## 2 Related Work

Klein, C. et al [14], in their study, reviewed the application of Artificial Intelligence algorithms in the field of tumour histopathology. The digitalization of histopathology slides at high magnification has raised the opportunity to extract important information due to image analysis. They highlight the usage of deep learning broadly for diagnosis, sub-typing, grading and prognosis of tumours as well as mentioning the potential challenges-economic, regulation and ethical issues.

Kosaraju, S. et al [15], introduced an architecture for slide-based histopathological image analysis-HipoMap. HipoMap converts a WSI to structure image-type representation and has shown a significant improvement in various metrics such as Area Under Curve (AUC) for classification of lung cancer and c-index in survival analysis with TCGA lung cancer data respectively.

For grading breast cancer into two stages-low/intermediate and high, Wetstein, S. et al [16], used a Multiple Instance Learning deep learning model architecture that utilized ResNet-34 as the backbone. The model showed good performance in the prediction of survival analysis. This model, however, cannot distinguish between low (grade 1) and intermediate (grade 2) tumours. In their study, the WSIs were graded by a single pathologist, so inconsistency between the grades can make it harder for the model to learn important patterns in the dataset. Grading from multiple pathologists could have diluted this problem.

Wang, D et al [17], developed a deep learning model for automated tumour region recognition for lung cancer pathology images. They extracted well-defined shaped and boundary features and found that 15 of them were significantly associated with patient survival outcomes in lung adenocarcinoma patients from the National Lung Screening Trial. The tissue extraction was done using a predicted tumour region heat map generated by their model. They obtained the lung WSI dataset from two independent sources - NLST^3^ and TCGA^4^.

WSISA, introduced by Zhu, X et al [18], is a framework for the prediction of survival using WSI in cancer patients. Previous methods struggle to work with WSI because of its huge size whereas WSISA aims to exploit the dataset by using adaptive sampling and clustering techniques. Hence, this framework can represent a significant advancement in the application of image-based prediction methods for cancer prediction. Testing the model on survival prediction of Glioma and non-small-cell lung cancer, demonstrated the promising application of this framework on other related tasks.

The WSI datasets are easier to generate due to the advancement of technology in the medical field. Modern slide scanners can rapidly digitize entire tissue slides at high resolution. These images are giga-pixelated and take up a lot of space. However, annotating these datasets manually takes a lot of time and requires high precision from the pathologist. Due to this, there is an abundance of WSI datasets that are not annotated. Self-supervised learning (SSL) methods can be used to train on these datasets to leverage the important features and then fine-tuned on annotated datasets for specific tasks. Fan, L. et al [19], employed two kinds of SSL methods as pretext tasks to pre-train a covnet-based model. Then they aggregated the features from multiple WSIs using consistency and contrastive losses to normalize slide-level features for patient-level survival prediction.

Many studies in this domain extract and process only image features and ignore the spatial information of patches from WSIs. SeTranSurv, proposed by Huang, Z et al [20], extracts patch features from WSIs through SSL and adaptively aggregates these features based on their spatial information and correlation between patches. Through experiments on large cancer datasets, the effectiveness of this model is demonstrated.

## 3 Methodology

### 3.1 Dataset

The dataset used for experiments was obtained from the TCGA repository, specifically from the breast cancer collection (BRCA) ^5^. A total of 100 patient samples were utilised in this study which collectively occupied over 100 GB of storage space. The dataset comprises 50 samples of patients who survived (censored) and the remaining were from deceased patients. The survival time ranges from 0 to 244 months, with a median survival time of 32.5 months.

Due to their large size, they cannot be fed into the network directly so the WSIs were divided into non-overlapping patches of size 256 × 256 pixels. Since many patches contained minimal or no tissue samples, they were discarded (Figure 1). This selection was based on the disk space they occupied. Those patches that contained little tissue samples were predominantly blank and more efficiently compressible, occupying less space. In particular, those patch images that were 80 KB or less in size were discarded. The patches were normalized and resized to 224 × 224 pixels before passing for training. Figure 2 shows the Kaplan-Meier curve for the selected samples used in this study.

**Figure 1:**
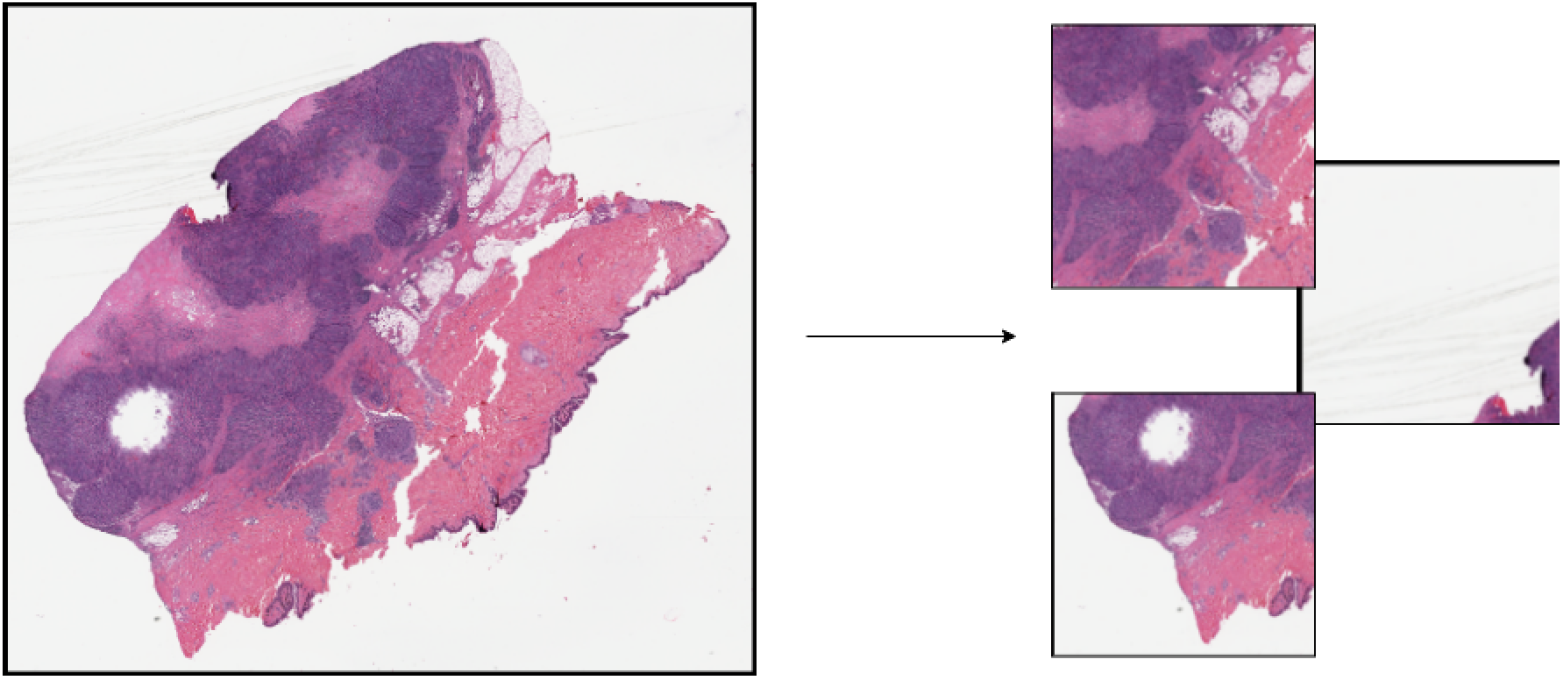
Whole Slide Images have high resolution. This one, extracted from breast tissue, has a resolution of 90440 × 80681 pixels. Dividing these images into patches of dimension 256 × 256 will result in some patches having minimal tissue samples.

**Figure 2:**
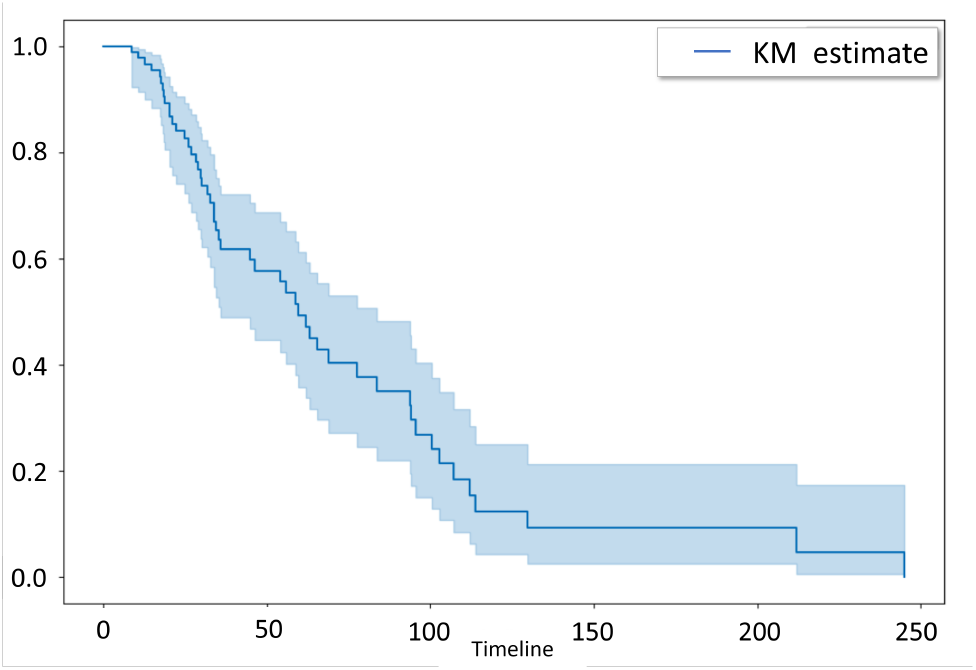
The Kaplan-Meier (KM) curve demonstrating the survival analysis for estimating survival probability over time.

### 3.2 Metrics

Negative partial log-likelihood [21] is used as a loss function for the experiments. Another popular method for evaluating a model for survival analysis is using the Concordance Index or C-index for short. It assesses a model by ranking how well a model can rank survival times of data samples based on their predicted risk score. It ranges from 0 to 1. A value of 0.5 indicates a model predicting random risk scores, while a value of 1 denotes that a model is predicting perfect risk scores. The lifeline library is used for the calculation of C-index. It is given by:

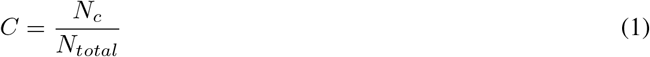

Where:

- *N*_*c*_ is the number of concordant pairs (where the predicted order matches the observed order).
- *N*_*total*_ is the total number of comparable pairs (pairs where at least one event occurred).

### 3.3 Pretrained Vision Encoder

Three publicly available pre-trained models have been used for generating the representation of WSIs for survival analysis. These vision encoders are frozen and will only be used to generate feature embedding for WSIs. Table 2 shows the model comparison for each of them. Since the number of embeddings generated by each model varies, the number of trainable parameter for entire model setup is also different.

**Table 1:** Most Common Cancers in 2020 (new cases) according to WHO.

| Type of Cancer | Number of New Cases |
| --- | --- |
| Breast | 2.26 million |
| Lung | 2.21 million |
| Colon and Rectum | 1.93 million |
| Prostate | 1.41 million |
| Skin (non-melanoma) | 1.20 million |
| Stomach | 1.09 million |

**Table 2:** Model Summary for each experiment. Note: The provided numbers pertain to the entire model setup, not just the vision encoder.

| Model | Parameters | Trainable Parameters | Encoder Embedding Size | Encoder Architecture |
| --- | --- | --- | --- | --- |
| UNI | 304,412,176 | 1,061,392 | 1024 | Transformer |
| Phikon | 86,712,592 | 913,936 | 768 | Transformer |
| ResNet18-TIAToolbox | 11,942,992 | 766,480 | 512 | ResNet18 |

#### UNI

UNI [22] is a general-purpose self-supervised model trained using more than 100 million images from 100k WSIs across 20 major organ types, making it the largest pretrained vision encoder for histopathology. The data used for training include some from private histology collection. Based on vision transformer architecture and trained using the DINOv2 [23] self-supervised training algorithm consisting of a mask image modelling objective and a self-distillation object as a pretext task, UNI demonstrates exceptional performance across 34 clinical tasks of varying diagnostic difficulty.

#### Phikon

Phikon [24] is a self-supervised learning model developed by Owikin for feature extraction from histopathology images. This transformer-based model is trained on 40 million pan-cancer tiles extracted from the TCGA dataset. It can be fine-tuned for specific downstream tasks like cancer classification across different subtypes.

#### ResNet18-TIAToolbox

Trained on kather100k^6^, also called NCT-CRC-HE, this ResNet model for tissue classification is made available by the TIAToolbox [25] which is a library for advanced tissue image analytics. The dataset contains 100k annotated patches from the colorectal tissue section. We are using a third-party implementation of the model for this study^7^.

### 3.4 Network Architecture

The entire network setup can be divided into three parts-Pretrained vision encoder, aggregator module and risk prediction head as shown in Figure 3. The input to the encoder is a 5-dimensional tensor (batch, number of patches from each wsi, channels, width, and height). The pretrained encoder allows the network to leverage transfer learning, utilizing knowledge learned from large-scale image datasets. The feature embeddings from the encoder are stacked together and passed into the aggregator function which is an attention layer that computes a weighted sum of these features, effectively aggregating the information while emphasizing the most informative patches. Finally, the output from the aggregator function is passed into the risk prediction head to generate the risk score for each wsi. The LSTM is configured with two layers and a hidden size of 128. It processes the sequence of attention-weighted features, capturing temporal dependencies and relationships among the patches. The input size of the LSTM layer changes as the size of feature embedding depends on the encoder used for its generation.

**Figure 3:**
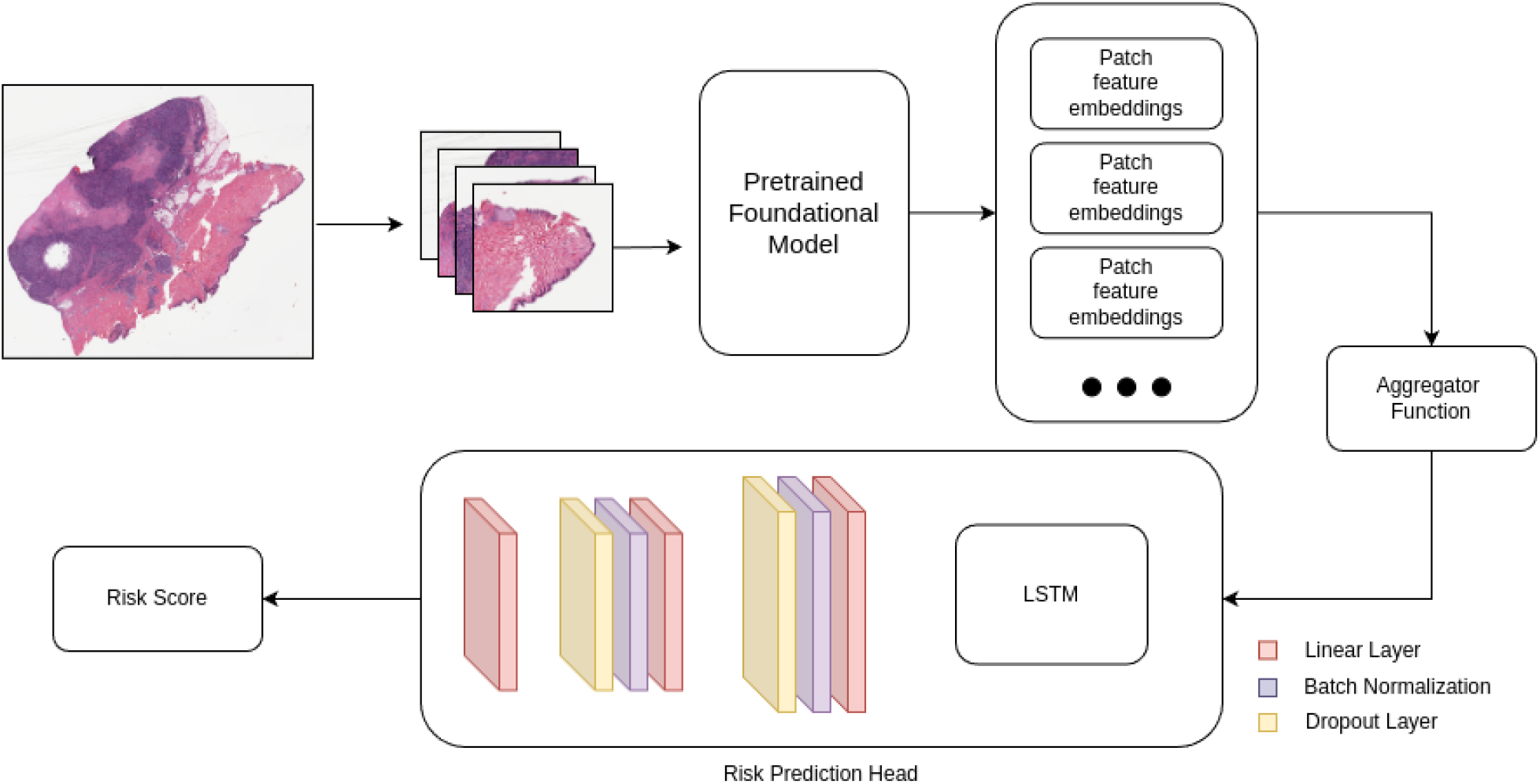
Medical Image Analysis Pipeline for Risk Prediction.

## 4 Experiments and Results

PyTorch is used for the experiment implementation. The system used for this research is configured with Ubuntu 23.10 as the operating system. It includes 16GB of DDR5 RAM, an AMD Ryzen™ 7 7840HS processor, and an Nvidia RTX 4060 GPU with 8GB of memory.

The number of patches resulting from the patch selection varies from wsi to wsi due to their variation in size. Out of these patches, 300 of them are randomly chosen to represent the wsi. The experiment is set up using a k-fold validation pipeline (k=4 for this study). It is a resampling method where the dataset is split into k subsets known as folds and trained k time using k-1 data for training and the rest for validation. After completing the training and validation for all folds, the results are averaged to provide an overall performance metric. This helps in assessing the model’s ability to generalize. Each of the experiments was performed using the same set of configurations. Table 3 shows the hyperparameter tuning values for the experiments.

**Table 3:**
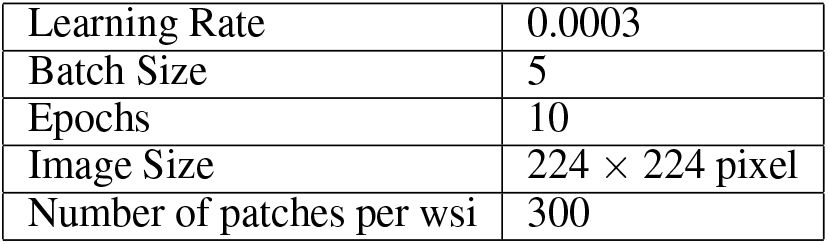
Hyperparameter Configuration.

Table 4 shows the performance of the experiments performed and evaluated using the C-index. The UNI model achieved a C-index of 0.5842 with a standard deviation of ±0.0366 whereas the Phikon model demonstrated slightly better performance with a C-index of 0.5872, however, it showed greater variability than the UNI as indicated by its high standard deviation making UNI model more reliable for survival analysis. The ResNet18 model trained on histopathological images performed worse than the transformer-based encoders with a C-index of 0.5689.

**Table 4:** Average Validation C-index and their standard deviation for each model for the 4-fold validation setup.

| Model | C-index |
| --- | --- |
| UNI | 0.5842 (± 0.0366) |
| Phikon | 0.5872 (± 0.1073) |
| ResNet18-TIAToolbox | 0.5689 (± 0.0139) |

## 5 Conclusion

This study evaluated the effectiveness of three different pre-trained vision encoders—UNI, Phikon, and ResNet18—on the task of survival analysis using Whole Slide Images (WSIs) from the breast cancer dataset in The Cancer Genome Atlas Program (TCGA). Our experiments demonstrated that transformer-based models-UNI and Phikon, outperformed the ResNet18-based model, with Phikon achieving the highest average C-index followed by UNI which performed a little worse than Phikon. However, UNI showed more consistency in performance and had less fluctuating C-index than Phikon.

These findings indicate the promising application of deep learning in computational pathology. C-index can range from 0 to 1, with 1 being the perfect prediction, indicating there remains significant room for further research. Future works could be conducted to explore different model’s applications in other datasets as well as to assess their generalizability.

## Data Availability

All data produced are available online at a private repository which will be made public. link- https://github.com/AsadNizami/Survival-Analysis

https://portal.gdc.cancer.gov/analysis_page?app=Downloads

## Footnotes

1 https://github.com/AsadNizami/Survival-Analysis

2 https://www.cancer.gov/about-cancer/understanding/statistics

3 https://biometry.nci.nih.gov/cdas/learn/nlst

4 https://portal.gdc.cancer.gov/

5 https://portal.gdc.cancer.gov/projects/TCGA-BRCA

6 https://tia-toolbox.readthedocs.io/en/latest/_autosummary/tiatoolbox.models.dataset.info.KatherPatchDataset.html

7 https://huggingface.co/1aurent/resnet18.tiatoolbox-kather100k

